# Clustering of Countries for COVID-19 Cases based on Disease Prevalence, Health Systems and Environmental Indicators

**DOI:** 10.1101/2021.02.15.21251762

**Authors:** Syeda Amna Rizvi, Muhammad Umair, Muhammad Aamir Cheema

## Abstract

The coronavirus has a high basic reproduction number (*R*0) and has caused the global COVID-19 pandemic. Governments are implementing lockdowns that are leading to economic fallout in many countries. Policy makers can take better decisions if provided with the indicators connected with the disease spread. This study is aimed to cluster the countries using social, economic, health and environmental related metrics affecting the disease spread so as to implement the policies to control the widespread of disease. Thus, countries with similar factors can take proactive steps to fight against the pandemic. The data is acquired for 79 countries and 18 different feature variables (the factors that are associated with COVID-19 spread) are selected. Pearson Product Moment Correlation Analysis is performed between all the feature variables with cumulative death cases and cumulative confirmed cases individually to get an insight of relation of these factors with the spread of COVID-19. Unsupervised k-means algorithm is used and the feature set includes economic, environmental indicators and disease prevalence along with COVID-19 variables. The learning model is able to group the countries into 4 clusters on the basis of relation with all 18 feature variables. We also present an analysis of correlation between the selected feature variables, and COVID-19 confirmed cases and deaths. Prevalence of underlying diseases shows strong correlation with COVID-19 whereas environmental health indicators are weakly correlated with COVID-19.

## 1. Introduction

Pandemics and epidemics can lead to a large number of fatalities in merely a few days. With the increase in population growth rate, the rate of infectious diseases is also growing. COVID-19 has caused 2,260,259 deaths around the world (World Health Organization estimate as of, Feb 4, 2021).

Pandemics lead to interference in economic development, resulting in shortfall of basic foods, inflation, decrease in Gross Domestic Product (GDP) and threat to lives. For example, a serious pandemic may reduce GDP by 3-4% [1]. The adverse effects of Influenza pandemic on the GDP are discussed in [2]. Due to the influenza pandemic, the businesses were severely affected mainly due to a decrease in demand and many businesses were closed resulting in an increase in unemployment [3]. The Ebola epidemic in 2014 had catastrophic effects on economy in several ways as it lead to depletion of agricultural production, inflation, higher unemployment rates and a decrease in trade, tourism, and investments [4].

World is facing a second wave of COVID-19 and new variants of COVID-19 have also appeared such as the UK variant of COVID-19 [5]. According to the studies and health experts, it is always helpful to know the factors associated with the transmission of disease [6]. These factors may include health system indicators, disease prevalence, and other variables which can indirectly cause the rapid spread of disease. The relation of these factors with various pandemics have been explored by many researchers in the past. A research study was conducted to cluster the countries on the basis of epidemic preparedness index in order to identify the countries’ readiness and the strategy to respond to such outbreaks [7]. As the ways of infection transmission holds significant importance, an analysis was performed on multiple ways of Varicella-Zoster virus transmission which gives insights related to the type of contact clusters highly involved in transmission [8]. There is a dire need to develop similar clustering analysis for COVID-19 as well, so that policy makers can take better decisions to mitigate its spread.

Machine Learning (ML) and Artificial Intelligence (AI) are commonly used to study the factors responsible for the epidemics outbreak. Such a system to overcome the tract of transmission of infectious diseases is developed by Agrebi *et al*. [9] which detects infected patients through classification using vital signs. There are only a few examples of clustering of countries for COVID-19. Carrillo-Larcois *et al*. [10] present an analysis of country wise variables for stratifying countries on the basis of COVID-19 confirmed cases leading to useful studies of the country profiles to better analyse the relationship between factors involved in the spread of disease. Another work by Farseev *et al*. [11] is the study of economic and health factors impacting the COVID-19 disease spread resulting in the formation of four country clusters. Siddiqui *et al*. [12] explore the relation of COVID-19 confirmed, suspected and death cases with temperature profiles. K-means is used to cluster different regions of China. Hubei and Hainan depict similar effects being at same temperature profiles. It identifies that temperature is not the only factor which affects COVID-19 spread. Another analysis by Imtyaz *et al*. [13] on the COVID-19 data depicts impacts of governments’ response to COVID-19. It concludes that age is the most significant factor relating to death cases. While lock-down is another significant factor in controlling COVID-19 confirmed cases [13].

To the best of our knowledge, this is the first work that finds the correlation of disease prevalence, and socioeconomic and environmental indicators with the spread of COVID-19 during the second wave. We perform an analysis on a data set of 79 countries and 18 feature variables both for the COVID-19 confirmed cases and COVID-19 deaths. Since the beginning of COVID-19, a number of studies have been published that perform clustering for COVID-19. Some papers discuss the relation of feature variables for a limited number of countries [14]. While some models fail to stratify countries for fatality rates [15]. In this paper, a detailed analysis finding similarities and differences among different groups of countries have also been provided. Existing articles only provide comparative analysis for a limited set of countries [16]. So, existing literature lacks discussion of refined and detailed approach towards the data analysis process for discovering pandemic situation all around the globe which can also be replicated for future pandemics.

In this paper, data from different sources – global health observatory, World Health Organization (WHO), World Bank and global health data exchange websites – is acquired for 79 countries. Description of these feature variables has been presented in Table 1. The factors selected for the analysis are clinically and medically related to COVID-19. Pearson Product Moment Correlation analysis is used to explore the most significant factors from health system indicators (PM2.5 exposure, unsafe sanitation, unsafe drinking water, air quality, sanitation and drinking water score), disease prevalence (tuberculosis, cardiovascular disease, respiratory infections, asthma, nutritional deficiencies) and socio-economic factors (GDP per capita, health expenditure per capita, alcohol consumption, smoking prevalence and life expectancy) that may indirectly affect rapid spread of COVID-19. Unsupervised K-Means algorithm is used to cluster the countries considering all the above mentioned factors. Four clusters are formed. China and India lies in one cluster with maximum number of COVID-19 cases. Cluster containing developed countries have relatively higher number of COVID cases. Asthma, Diabetes mellitus, respiratory infections, nutritional deficiencies and tuberculosis show strong correlation with COVID-19. A weak correlation exists for alcohol consumption, environmental health index and life expectancy. Smoking prevalence shows negative association with COVID-19. This negative correlation may be due to presence of nicotine receptors in smokers which reduce the likelihood of getting infected with COVID-19 [17]. This detailed analysis of the relation between feature variables results in determining the potential indicators responsible for the spread of COVID-19. This paper also presents a novel analysis of the factors that are responsible for formation of clusters using the count of COVID-19 confirmed and COVID-19 death cases. This leads to useful insights related to a country’s strategies that are impacting COVID-19 prevalence.

**Table 1.** Description of Feature Variables

| Notions and Data Sources | Variable Name | Description |
| --- | --- | --- |
| COVID-19 cases<br>**Source:** World Health Organization (WHO COVID-19 Dashboard) [18] | Cum_confirm cases | Cumulative confirmed COVID-19 cases. |
|  | Cum_deaths | Cumulative COVID-19 death cases. |
| Socio-economic Indicators<br>**Source:** World Bank[19], WHO's global health observatory data repository [20] | GDP_per_capita | Gross domestic product per capita, is a proportion of a country's financial yield represented by the number of individuals. |
|  | Health Exp | Health expenditure per capita is the average amount that country is devoting for health services for an individual. |
|  | Alcohol Consumption | Alcohol consumption rate per capita by an age group of 15+. |
|  | Smoking Prevalence | Pervasiveness of smoking is the level of people over the age of 15 who presently smoke any tobacco item on a day by day or non-regular schedule. |
|  | Life Expectancy | It is the anticipated years a baby would live if accepted examples of mortality at childbirth were to remain the equivalent for the duration of its life. |
| Disease Prevalence<br>**Source:** Global Burden of Disease Collaborative Network, Global Burden of Disease Study 2017 (GBD 2017) Results. Seattle, United States: Institute for Health Metrics and Evaluation (IHME), 2018 [21] | Tuberculosis cases | Prevalence in terms of cases of tuberculosis disease. |
|  | Cardiovascular Disease | Prevalence in terms of number of cases due to known cardiovascular causes such as ischaemic heart disease, stroke e.t.c. |
|  | Diabetes Mellitus | Prevalence in terms of cases of diabetes mellitus. |
|  | Respiratory Infections | Prevalence in terms of cases of Respiratory infections such as pneumonia and bronchitis. |
|  | Asthma | Prevalence in terms of cases of chronic lung disease asthma. |
|  | Nutritional deficiencies | Prevalence in terms of cases of nourishing inadequacies including protein-energy unhealthiness, lack of iodine, nutrient A insufficiency, iron inadequacy, and other health insufficiencies. |
| Environmental Performance Indicators<br>**Source:** Environmental Performance Index, 2020 [22]. | PM2.5 exposure (PMD) | It is the indicator of number of people who have lost life years per 100,000 people due to exposure to fine air particulate matter smaller than 2.5 micrometers. |
|  | Environmental Performance Index (EPI) | It is a score assigned from 1-100, on the basis of how close countries are to set up health environmental targets |
|  | Environmental Health (HLT) | It gives the environmental health score of a country. |
|  | Air Quality (AIR) | Country score on the basis of effects of air contamination. |
|  | Household Solid Fuels (HAD) | Indicator of AIR issue category which gives score on the basis of lives lost due to use of household solid fuels. |
|  | Sanitation and Drinking Water (H2O) | Country score on basis of how well nations shield human well-being from natural dangers on two pointers: UWD and USD. |
|  | Unsafe Water for Drinking (UWD) | Country score on basis of people who have lost life years per 100,000 people due to insufficient proper drinking water facilities. |

The rest of the paper is organized as follows: Section II details existing studies. Section III presents the methodology being followed to cluster the countries. Section IV and V present analysis of clustering results for COVID-19 confirmed cases and COVID-19 death cases, respectively. Section VI presents choropleth maps of clusters being formed. Finally, Section VII concludes the paper.

## 2. Related Work

Since the beginning of COVID-19, researchers and clinicians are trying to mitigate the spread of COVID-19. Carrillo *et al*. [10], uses unsupervised machine learning to classify 155 countries that share similar COVID-19 profile. Clustering is performed for COVID-19 confirmed cases. Disease prevalence, male population, air quality index, socio-economic metrics and health system indicators are used as feature variables. The clusters formed give insights about similarities and differences among countries in-terms of impact by COVID-19. This model fails to stratify countries on the bases of COVID-19 fatality rate. Another work by Farseev *et al*. [11], covers similar economic and health factors for COVID-19 spread. The study unveils significant relationships between COVID-19 and other national statistics. It identifies four clusters on the basis of country’s economy and health system indicators. Stojkoski *et al*. [23], present an analysis on the socio-economic determinants of COVID-19. It determines the socio-economic, health care, demographic and environmental factors which are more or less involved in the spread of COVID-19. A stream of work by Zarikas *et al*. [24], is the introduction of a clustering algorithm especially designed for the clustering of countries based on the COVID-19 active cases, active cases per population and per area following the concept of hierarchical analysis. The results lead to the analysis that countries which face similar impact of COVID-19, possess same societal, economical and other factors.

Aungkulanon *et al*. [25] perform clustering of different regions of Thailand based on financial conditions and mortality differentials. Cluster examinations uncover super-locale (groups of already merged districts) which are prevalently urban and have low all-cause normalize mortality proportion yet a high colorectal disease explicit death rate. Deaths caused by liver malignant growth, diabetes, and renal sicknesses are regular in low financial super-regions. Efficacy of digital tools is playing a significant role for surveillance of information streams, search designs, and the related advanced socio-economics at a very large scale. Such an adequate tool is used by WHO, originally cautioned of a secretive new respiratory infection in Wuhan, China. A group of specialists caught clues about the episode from online press reports and delivered their discoveries in a real time framework called HealthMap [26]. A study is conducted by Malav *et al*. [27] to forecast coronary illness using K-means and artificial neural networks. Only 14 instances of heart diseases are considered and this combined approach lead to a system with very high exactness rate. Another work by Singh *et al*. [28], forecasts heart diseases by grouping the data and then classifying using K-means and Logistic Classifier with high accuracy.

Clustering of countries to analyze different variables associated with a pandemic has been a topic of interest for researchers. Isikhan *et al*. [29], cluster the countries on the basis of causes of deaths, health profiles and risk factors. Unsupervised KMeans is used and clusters are analyzed on some financial and socio-demographic pointers. The findings point out that climate and ethnicity are more significant for clustering rather than socioeconomic factors. Grein *et al*. [30] analyzes different variables related to technology, economy, health, culture and life quality and observes the effect of corruption performance index over the course of 3 years for 39 countries. Hierarchical clustering is applied to study the cluster membership. The findings show that there is a strong relation between the corruption and GDP. Nastu *et al*. [31], clusters the countries in to two groups (economically developed and economically underdeveloped countries) on the the basis of economic aspects considering 12 factors which prove to be helpful for analyzing the economic progress of countries. Contribution of Anderson *et al*. [32] is the cluster analysis of non-OECD (Organisation for Economic Co-operation and Development) countries classifying them into groups of “chronically deprived”, “good performers”, and “others” group on the basis of seven quality of life indicators. Findings reveal well separated clusters on the basis of four strong indicators among seven that are GDP per capita, child mortality, fertility and under-nourishment. Tosto *et al*. [33], applied K-Means grouping on 3,502 patients of Alzheimer’s disease with longitudinal appraisals from the National Alzheimer’s Coordinating Center’s information base, incorporating 394 patients providing neuropathological information. It reveals high extrapyramidal burden while clusters show significantly greater number of patients diagnosed with dementia with Lewy bodies.

Kumar *et al*. [34] propose a combination of K-means and Support Vector Machine (SVM) to forecast the confirmed COVID-19 cases and to analyze the recovery rate, taking in to account the closely related factors involved in the increasing COVID-19 confirmed cases. An efficient COVID-19 predictor utilizing IoT devices for acquisition of data is also developed. Research conducted by Hu *et al*. [35] is the efficient model designed to label the CT images of COVID and Non-COVID patients showing precisely the location of any contusion if present which proves to be very beneficial for patient recovery. Brunese *et al*. [36] conducted a research to classify the X-ray images with COVID-19 disease and pulmonary disease and to notify the presence of COVID-19 disease in the regions that might be of interest for medical interpretability. Deep learning is used for efficient and cost-effective COVID-19 disease detection.

It has been discovered by many researchers that COVID-19 spread is widely affected by weather profiles. Malki *et al*. [37] find the relation between COVID-19 mortality rate and weather factors, using linear machine learning models such as Linear Regression and Least Angle Regression etc. Strongly related features of weather are deduced from correlation analysis and it is inferred from the outcome that both temperature and humidity are significant highlights for anticipating COVID-19 death rate. Study conducted by Sahin *et al*. [38] inspects the relationship between climate factors and COVID-19 disease spread in different cities of Turkey. The investigations are conducted using Spearman’s relationship coefficients and the outcomes demonstrate that significant relationships exist for population, last fortnight wind speed and temperature. Rosario *et al*. [39] assess the connection between climate factors (temperature, mugginess, sunlight based radiation, wind speed, and precipitation) and COVID-19 contamination in the State of Rio de Janeiro, Brazil. High temperature and wind speed are found to be the significant components affecting the spread of COVID-19.

There are some major differences between our paper and the above mentioned articles. Firstly, we have clustered the countries for both COVID-19 confirmed cases and COVID-19 death cases involving all the 18 feature variables. Some of the above discussed articles performs clustering only for a single notion [10]. Secondly, we have cumulatively studied the impact of disease prevalence along with other variables affecting the spread of COVID-19 in a wide set of countries. While existing studies only provide analysis on a limited set of countries [14]. Finally, our paper is the first work that clusters a wide set of countries using COVID-19 data in the second wave.

## 3. Methodology

In this section, we provide the details of our methodology to cluster the countries on the basis of selected feature variables.

### 3.1. Data Sources

Dataset consists of 18 feature variables based on four notions mentioned in Table 1. Socio-economic indicators comprise of GDP per capita, health expenditure per capita, alcohol consumption, smoking prevalence and life expectancy. Disease prevalence rates in selected countries include tuberculosis, cardiovascular disease, respiratory infections, asthma and nutritional deficiencies. Health system indicators entail six indicators that are based on Environmental Performance Index (EPI) which gives an information driven outline of the condition of support ability around the globe. It has 11 issue classes and 32 performance pointers. These markers provide a measure of exposure of the general public to environmental pollutants. This paper focuses on only 2 issue categories: Air Quality, and Sanitation and Drinking Water. These indicators are identified with the COVID-19 pandemic, both from a clinical and general well-being view-point. Diabetes mellitus adds to the severity of COVID-19 patients [40]. Same is the case with the disease prevalence of respiratory infections, tuberculosis and cardiovascular disease [41]. In this paper, we also discuss the financial status and well-being of countries, which affect the likelihood of an individual to adopt the preventive measures.

Next step in data pre-processing is feature scaling, as some of the variables such as GDP per capita, asthma prevalence, respiratory infections and nutritional deficiencies have large variance and it makes the model learn from the most dominate features. Standardization is used to centralize the data by removing the mean value of each feature and then scale it by dividing (non-constant) features by their standard deviation. After standardizing data, the mean will be zero and the standard deviation will be equal to one [42]. Standard Scalar method available in scikit-learn purely centralizes the data using the formula given in Eq. (1) for every value *x*_*i*_ in a set of feature values *X*. Hence features are close to normal distribution.

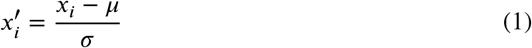

where *x*_*i*_ is the raw feature value (i.e., before standardization), and *µ* and *σ* are the mean and standard deviation of *X*, respectively. 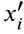 is the standard score and represents the number of standard deviations by which *x*_i_ is above (if 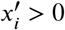) or below (if 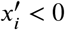) the mean of *X*.

### 3.2. Number of Clusters

The next step is to find the optimum number of clusters using Elbow method. The goal is to find an appropriate value of K (the parameter to be fed to the K-Means algorithm) for which total intra-cluster variation is minimum. To find K (Optimal number of clusters), the sum of squared distances (usually Euclidean distance) of samples with the nearest cluster center is calculated. As the value of K increases, average distortion – the average of squared distances from the cluster centers – decreases and each cluster instance becomes closer to the respective centroid. The value of K at which inertia decreases is the elbow (bend), the indicator of optimal number of clusters. As shown in Fig. 1, the bend indicates that optimal value for K number of clusters is 4. Now to cluster the countries on the basis of socio-economic, disease prevalence and other health indicators, all the components are fed to the K-Means algorithm with the pre-defined number of clusters, deduced from elbow method.

**Figure 1:**
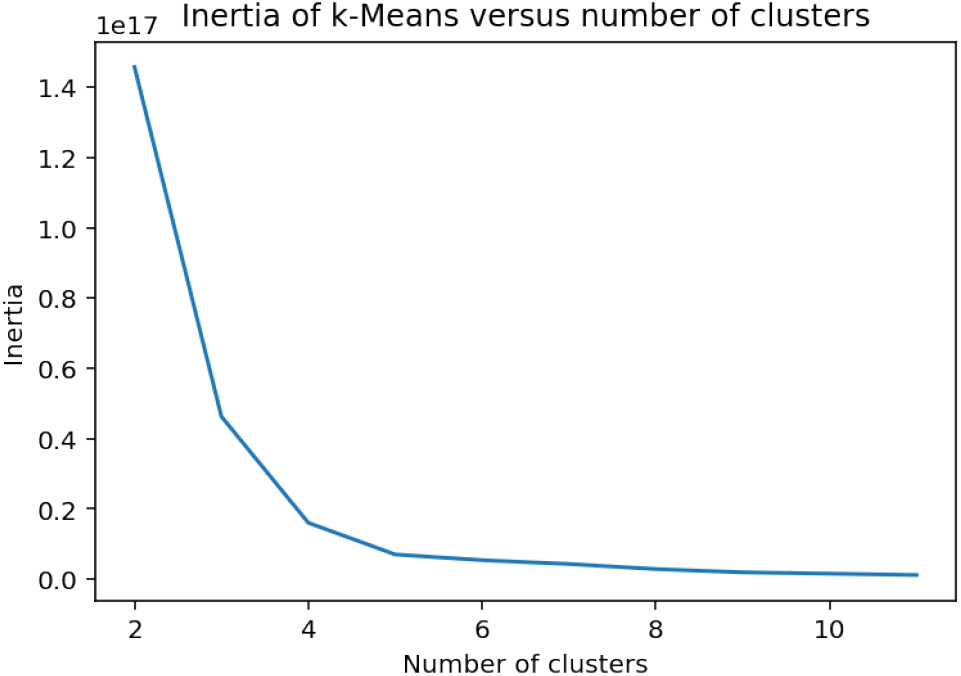
Optimal Number of Clusters using Elbow Method

### 3.3. Clustering Technique

K-Means is the most commonly used clustering algorithm because of its simplicity and effectiveness. It forms clusters with high intra-cluster similarity and low inter-cluster similarity, i.e., samples within the same cluster are very similar whereas the distances between samples from different clusters are large. In our work, we have used Centroid-based Partitional clustering. The number of cluster centers are pre-defined and on each iteration, the mean of cluster centers is updated on the basis of reassigned data points at a minimum distance from respective cluster centers. Each country has feature variables encompassing socio-economic indicators, disease prevalence and health system indicators. Membership is assigned to each country’s feature variables such that initially the K cluster centers are defined and each country’s data-point is reassigned a corresponding cluster center on the basis of minimum distance from that cluster center and thus the new cluster mean is updated. As a result, feature variables depicting similar behaviour result in stratification of countries which is beneficial for deriving the relationships between different factors that lead to cluster membership. The clusters of the countries depict what aspects are leading to higher number of COVID-19 confirmed cases and COVID-19 deaths, leading to evaluation of countries’ strategies. Hence, better decisions can be made to mitigate the spread of pandemic.

## 4. Analysis for COVID-19 Confirmed Cases

In this section, we first analyse the correlation of feature variables for COVID-19 confirmed cases. Then, we present the results of clustering for COVID-19 confirmed cases and analyse each individual cluster.

### 4.1. Correlation of Features Variables

Correlation matrix is used to find the relationship between two variables. Pearson Correlation Coefficient (derived from standard score of feature variables) is used to calculate the strength of this relationship between two quantitative variables *X* and *Y* (each containing *n* values) by using the formula given in equation (2):

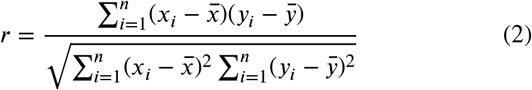

where *x*_*i*_ (resp. *y*_*i*_) is the *ith* value of *X* (resp. *Y*) and 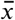 (resp. 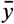) is the mean of *X* (resp. *Y*). If the Pearson Correlation is +1, it indicates presence of strong positive correlation, i.e., if value of one variable increases the other one also increases and vice versa. If value is near −1 it indicates a strong negative correlation, i.e., if value of one variable increases the other one decreases and vice versa. As shown in the Fig. 2 there is positive as well as negative association between cumulative confirmed COVID-19 cases and other 17 features: GDP per capita, environmental performance index (EPI), HLT (environmental health), air quality, fine particulate matter (PM_2.5_), H_2_O, asthma, cardiovascular disease, diabetes mellitus, nutritional deficiencies, respiratory infections, health expenditure per capita, tuberculosis, rate of alcohol consumption, life expectancy at birth. A high negative association exists between COVID-19 confirmed cases and smoking prevalence. Although the research is not conclusive yet, this negative correlation may be due to presence of nicotine receptors in smokers which reduce the likelihood of getting infected with COVID-19 [17].

**Figure 2:**
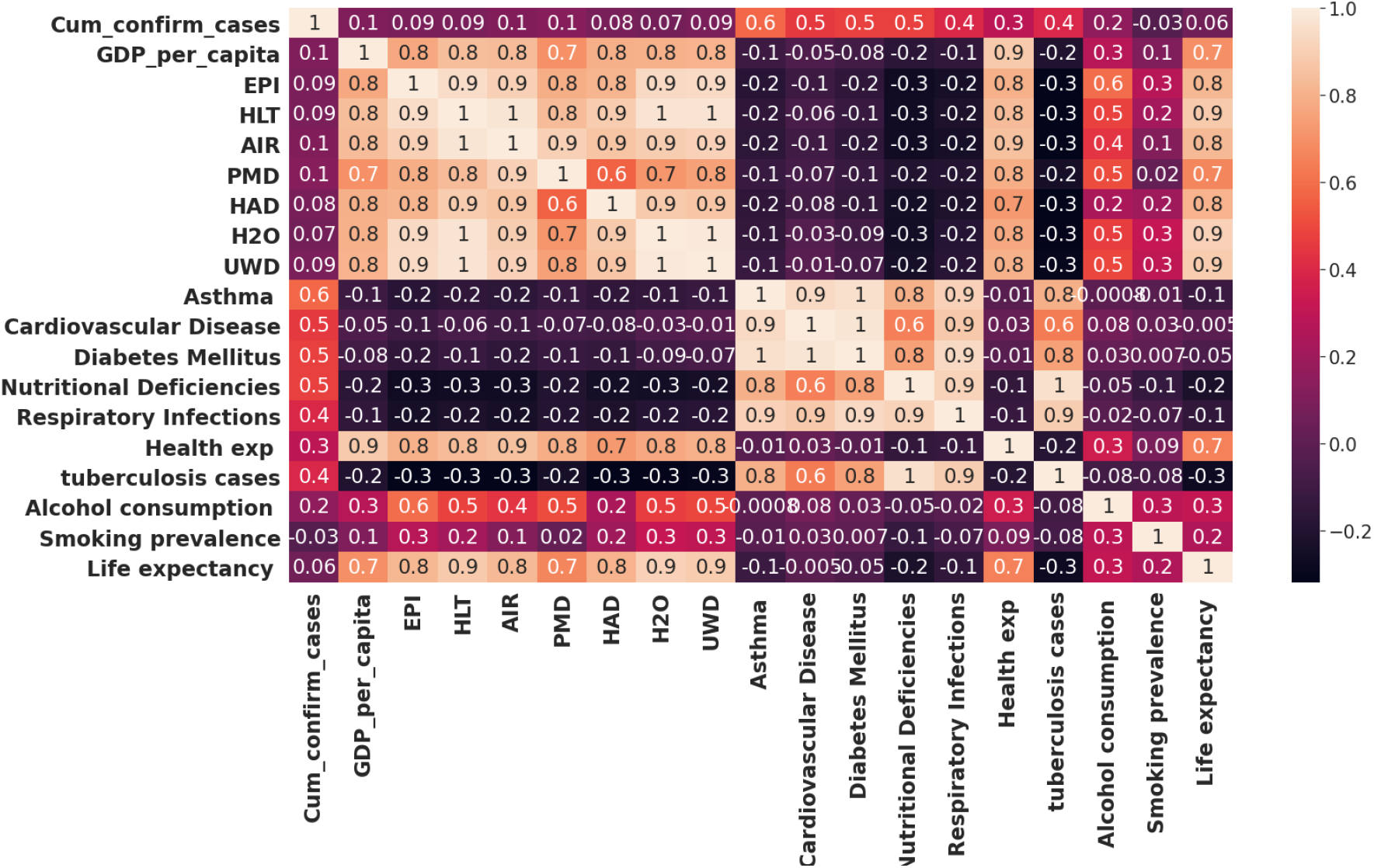
Pearson Correlation Heatmap for COVID-19 Confirmed Cases

It can be interpreted that high positive correlation exists between cumulative COVID-19 confirmed cases and asthma **(0.6)**, cardiovascular diseases (**0.5**), diabetes mellitus (**0.4**), respiratory infections **(0.5)**, nutritional deficiencies **(0.5)** and tuberculosis **(0.4)**. Moderate correlation exists between cumulative confirmed cases and health expenditure **(0.3)**. Correlation matrix shows that very weak association exists for alcohol consumption **(0.2)**, environmental health (HLT) (**0.1**), sanitation and drinking water (H_2_O) (**0.1**), GDP per capita **(0.1)**, fine particulate matter (**0.1**), air quality (**0.1**), environmental performance index (**0.1**) and life expectancy (**0.1**).

### 4.2. Cluster Analysis

#### 4.2.1. Analysis of Cluster 1

Cluster 1 comprises of 33 countries as shown in Table 2a. This cluster contains developed as well as developing countries. It has third highest correlation of cumulative confirmed cases (**-0.065**), inline with disease prevalence of respiratory infections (**-0.194**), tuberculosis(**-0.250**), nutritional deficiencies (**-0.189**) and diabetes mellitus (**-0.212**). Cardiovascular disease (**-0.189**) and asthma (**-0.246**) have least association as compared to other clusters. Health expenditure (**-0.412**) has a relatively higher correlation than that of cluster 4 that has least number of COVID-19 confirmed cases. According to the association results, nutritional deficiencies, respiratory infections and tuberculosis cases show strong correlation with the COVID-19 confirmed cases. These countries have very low child and adult mortality rate due to environmental risks according to WHO [43]. The results in Table 3 also indicates that this cluster has second highest life expectancy rate (**0.11**). It has second highest health performance index percentage (**-0.10**) and environmental performance indicator (**-0.079**).

**Table 2.** Clusters Produced based on COVID-19 Cases

| CLUSTER 1 |
| --- |
| Albania, Algeria, Argentina, Armenia, Bahrain, Belarus, Bosnia and Herzegovina, Brazil, Bulgaria, Chile, Colombia, Costa Rica, Croatia, Dominican Republic, Ecuador, Hungary, Iran (Islamic Republic of), Iraq, Kazakhstan, Kuwait, Malaysia, Mexico, Oman, Panama, Poland, Qatar, Romania, Russian Federation, Saudi Arabia, Serbia, Turkey, Ukraine, United Arab Emirates |

(b) Cluster No 2
| CLUSTER 2 |
| --- |
| Austria, Belgium, Canada, Denmark, France, Germany, Iceland, Ireland, Israel, Italy, Japan, Luxembourg, Netherlands, Norway, Portugal, Singapore, Spain, Sweden, Switzerland, United States of America, United Kingdom. |

(c) Cluster No 3
| CLUSTER 3 |
| --- |
| China, India |

(d) Cluster No 4
| CLUSTER 4 |
| --- |
| Afghanistan, Azerbaijan, Bangladesh, Bolivia, Djibouti, Ethiopia, Egypt, Ghana, Guatemala, Honduras, Indonesia, Madagascar, Mauritania, Morocco, Nepal, Nigeria, Pakistan, Philippines, Senegal, South Africa, Sudan, Uzbekistan, Zambia |

**Table 3.** Cluster Mean of Variables for COVID-19 Confirmed Cases

| Feature Variables | Cluster1 | Cluster2 | Cluster3 | Cluster4 |
| --- | --- | --- | --- | --- |
| Cum_confirm_cases | -0.065 | 0.241 | 1.906 | -0.292 |
| GDP_per_capita | -0.300 | 1.367 | -0.611 | -0.763 |
| EPI | -0.079 | 1.371 | -1.187 | -1.035 |
| HLT | -0.100 | 1.451 | -0.957 | -1.097 |
| AIR | -0.1812 | 1.475 | -1.184 | -0.983 |
| PMD | -0.351 | 1.392 | -0.996 | -0.680 |
| HAD | 0.043 | 1.220 | -1.007 | -1.088 |
| H<sub>2</sub>O | -0.000165 | 1.344 | -0.688 | -1.167 |
| UWD | -0.090 | 1.412 | -0.633 | -1.105 |
| Asthma | -0.246 | -0.084 | 5.277 | -0.027 |
| Cardiovascular | -0.189 | -0.0219 | 5.494 | -0.185 |
| Diabetes Mellitus | -0.212 | -0.082 | 5.714 | -0.116 |
| Nutritional Def. | -0.189 | -0.218 | 5.172 | 0.021 |
| Respiratory Infect. | -0.194 | -0.208 | 6.043 | -0.056 |
| Health exp | -0.412 | 1.441 | -0.615 | -0.670 |
| tuberculosis cases | -0.250 | -0.279 | 4.956 | 0.183 |
| Alcohol consumption | 0.128 | 0.578 | 0.046 | -0.715 |
| Smoking prevalence | 0.203 | 0.142 | -0.329 | -0.393 |
| Life expectancy | 0.118 | 1.122 | -0.400 | -1.160 |

#### 4.2.2. Analysis of Cluster 2

Cluster 2 in Table 2b comprises of 21 countries and this cluster contains most of the developed countries such as Norway, Ireland, Germany, Iceland and Singapore etc. This cluster has the highest GDP per capita (**1.367**) and EPI (**1.371**). Similarly environmental health index (HLT) and air quality have highest correlation **1.451** and **1.475**, respectively. It indicates a stable environmental health as compare to other clusters, but this cluster is more exposed to unsafe drinking water and PM_2.5_ fine particulate matter which can lead to short-term health effects such as sneezing, coughing and shortness of breath which are the symptoms similar to COVID-19. This cluster shows highest cluster mean (**1.122**) of life expectancy indicating the health stability and better health facilities in these counties.

It has second highest number of COVID-19 confirmed cases (**0.241**), as well as cardiovascular disease (**-0.0219**) and diabetes mellitus (**-0.082**), which are positively correlated with the confirmed COVID-19 cases. Asthma prevalence (**-0.084**) is third highest while nutritional deficiencies (**-0.218**), tuberculosis cases (**-0.279**) and respiratory infections (**-0.208**) have least mean as compared to other clusters. On contrary, some factors which are highly correlated with COVID-19 confirmed cases such as respiratory infections, nutritional deficiencies and tuberculosis cases have least mean values. Reason can be highest health expenditure (**1.441**), leading to better health capacities and better living.Another reason of second highest number of confirmed cases is better testing capacity resulting in more diagnosed cases as the countries involved have high GDP per capita and health expenditure.

#### 4.2.3. Analysis of Cluster 3

This cluster comprises of only two countries: China and India as shown in Table 2c. It has highest mean of COVID-19 confirmed cases (**1.906**). Asthma (**5.277**), cardiovascular disease (**5.494**), diabetes mellitus (**5.714**), nutritional deficiencies (**5.17**), respiratory infections (**6.043**) and tuberculosis cases (**4.956**) are significantly correlated with COVID-19 confirmed cases and has highest mean as compare to other clusters. It is in support to the assumption that the countries with high disease prevalence and having higher nutritional deficiencies are more likely to have greater number of COVID-19 confirmed cases. According to the recent studies, China’s population is growing old and chronic illnesses are spreading at a faster rate. Reasons are the lack of regular physical activity, smoking and unhealthy diet leading to obesity and higher death rates due to cardiovascular diseases and diabetes. Higher death rates have been reported in China due to ischemic heart diseases, chronic obstructive pulmonary disease and diabetes [44] and hence, making the patients more vulnerable to COVID-19 pandemic. Several studies indicate that poor dietary habits lead to nutritional deficiencies, hence, causing diseases such as cardiovascular complications, diabetes and obesity [45]. It supports the results that nutritional deficiencies (mainly Vitamin D) are considered as a risk for COVID-19 [46]. Similarly Vitamin C which is considered as an immunity booster, its deficiency also leads to higher risks of getting affected. India’s top leading causes of deaths include cardiovascular disease, respiratory diseases and tuberculosis hence making the community more prune to infections. The speedy transition from rural to urban areas is one of the reasons of increasing number of cardiovascular disease prevalence [47]. Other factors such as air quality (**-1.184**) and GDP per capita (**-0.611**) have third highest cluster means. PM_2.5_ fine particulate matter (**-0.996**) shows least cluster means. Air quality index of India and China are very low (**13.4 and 27.1** respectively), which triggers asthma, shortness of breath and cardiovascular problems.

#### 4.2.4. Analysis of Cluster 4

This cluster consists of least developed countries such as Afghanistan, Nepal, Sudan, Djibouti, Ethiopia, Mauritania, Madagascar, Zambia and Senegal as well as developing countries. as shown in Table 2d. This cluster comprises of 23 countries showing least cluster means of COVID-19 confirmed cases (**-0.292**). Other disease prevalence such as asthma (**-0.027**), diabetes mellitus (**-0.116**), nutritional deficiencies (**0.021**), respiratory infections (**-0.056**) and tuberculosis cases (**0.183**) show second highest cluster mean values. While, life expectancy (**-1.160**), cardiovascular disease (**-0.185**), alcohol consumption (**-0.715**) and health expenditure (**-0.670**) have least cluster means, supporting the assumption of few number of COVID-19 cases. This clusters involves a larger number of countries that have high child and adult mortality rate due to environmental factors, causing disease burden according to World Health Organization [43].

Disease prevalence is highly correlated with COVID-19 confirmed cases. Results show strong associated factors with COVID-19 cases i.e. high rates of asthma prevalence, diabetes mellitus, nutritional deficiencies, tuberculosis and respiratory infections. Whereas, there are least percentages of COVID-19 confirmed cases in this cluster. Despite the fact that this clusters involves a larger number of countries that have high child and adult mortality rate due to environmental factors causing disease burden according to WHO [43].

## 5. Analysis for COVID-19 Deaths

### 5.1. Correlation of Feature Variables

Correlation matrix in Fig. 3 shows significant correlation (**0.5**) of COVID-19 death cases with asthma prevalence. The significant positive correlation of COVID-19 confirmed cases are found with asthma (**0.5**), cardiovascular disease prevalence (**0.4**), diabetes mellitus (**0.4**), nutritional deficiencies (**0.4**), respiratory infections (**0.3**), tuberculosis cases (**0.3**) and health expenditure (**0.3**). There are less significant correlations with other factors such as PM_2.5_ fine particulate matter (**0.2**), alcohol consumption rate (**0.2**), life expectancy (**0.09**), GDP per capita (**0.08**), Environmental Performance Index (**0.1**), air quality (**0.1**), and unsafe sanitation and drinking water (**0.1**).

**Figure 3:**
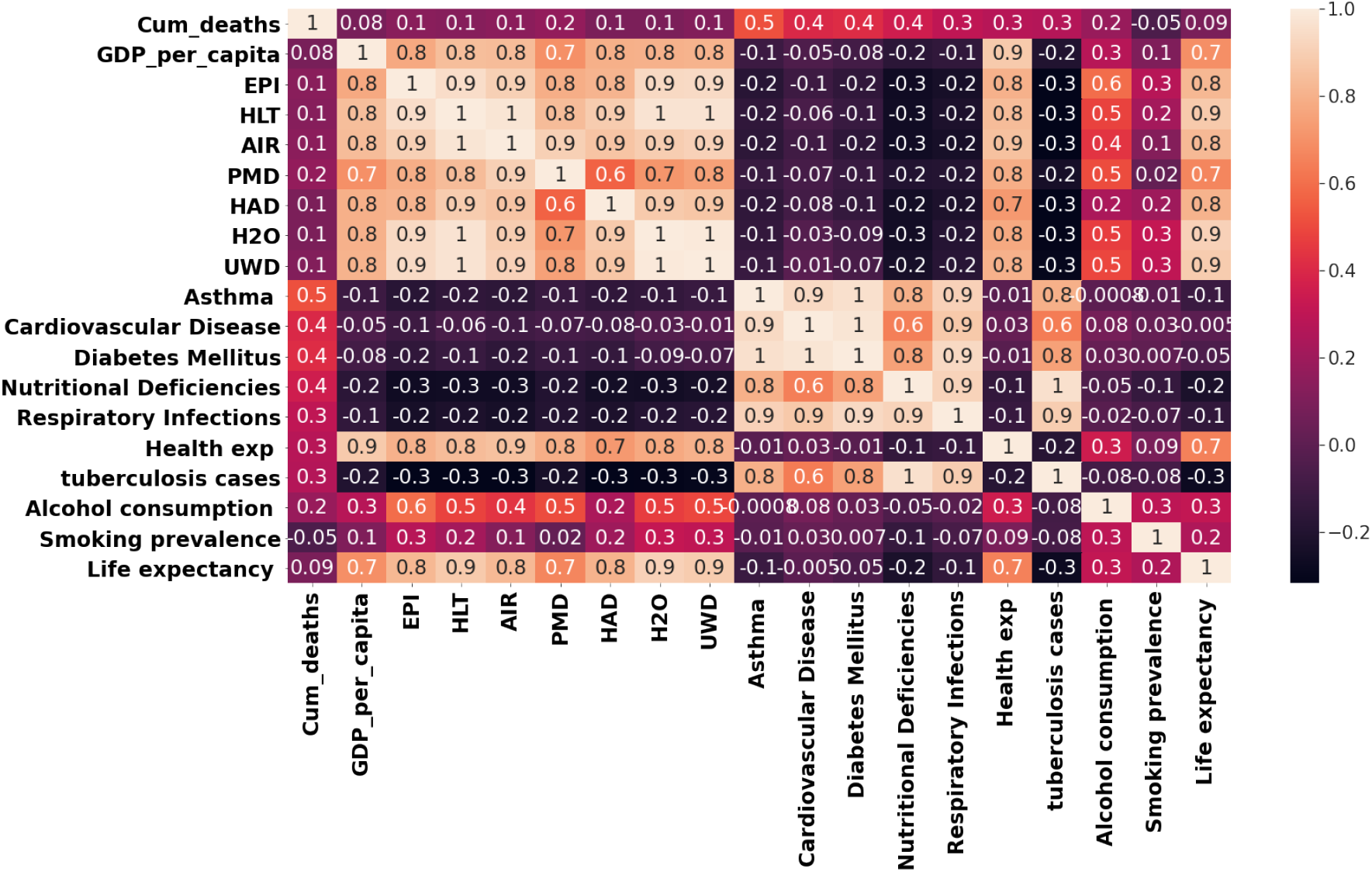
Pearson Correlation Heatmap for COVID-19 Death Cases

It can be deduced that the countries with asthma, diabetes mellitus, nutritional deficiencies, respiratory infections and cardiovascular disease prevalence are likely to have more number of COVID-19 deaths as compare to other countries. It can also be inferred that countries spending in healthcare are likely to report more death cases and confirmed cases due to the fact that they are keeping better track of the cases, thus reporting large number of affected cases and death rates accurately. It cannot be said that high health expenditure of a country is negatively correlated with the case fatality rate. Health capacity factor is a more suitable indicator which is likely to have strong sensitive correlation with COVID-19 mortality rate. According to a recent study, countries with greater number of nurses and midwives per one thousand people, the numbers of physicians per one thousand people and the numbers of hospital beds per one thousand people are likely to have fewer deaths per confirmed cases [48]. The studies have shown that people suffering from respiratory illnesses and asthma are at higher risk of COVID-19, thus leading to pneumonia or acute respiratory disease [49] and the severe complications that may lead to death. Similarly, people with cardiovascular disease, diabetes mellitus and other complications are at higher risk of facing severe complications due to COVID-19. Recent studies show that countries with higher levels of PM_2.5_ exposure are likely to have higher number of confirmed cases and death cases [50]. Correlation matrix also indicates that PM_2.5_ particulate matter is highly correlated with COVID-19 death cases.

Similar clusters (as produced for COVID-19 confirmed cases) are produced as a result of applying K-Means to cumulative death cases as shown in Table 2. Disease prevalence is strongly correlated to COVID-19 death cases. Further, the clusters formed for the evaluation of COVID-19 death cases are analyzed considering the cluster means of each variable.

### 5.2. Cluster Analysis

#### 5.2.1. Analysis of Cluster 1

The countries in cluster 1 are shown in Table 2b. It has second highest cluster mean of COVID-19 confirmed deaths (**0.241**). Cluster means show highest values of PM_2.5_ level (**1.392**) and have higher COVID-19 death cases as exposure to air pollutants rise the risks of death due to COVID-19. Other factors such as cardiovascular disease prevalence and diabetes mellitus have second highest cluster mean (**-0.0219, −0.082**). Most of the countries in this cluster have diabetes and heart diseases as leading causes of deaths. This cluster have least mean of nutritional deficiencies (**-0.218**) although these countries also have dietary risks as one of the top leading factor and has highest mean of health expenditure (**1.441**) and highest life expectancy (**1.122**). Countries like France, Germany, Italy and United Kingdom have excellent healthcare systems and have good ranking globally from which it is assumed that this set of countries have better testing facilities.

#### 5.2.2. Analysis of Cluster 2

The cluster 2 (shown in Table 2a) has third highest mean(**-0.0000306**) of COVID-19 death cases as mentioned in Table 4 and third highest mean of COVID-19 confirmed cases. Correlation matrix shows strong correlation of COVID-19 death cases with nutritional deficiencies, respiratory infections, tuberculosis and asthma, having third highest cluster means: **-0.1891, −0.194**,**-0.250 and −0.246** respectively. Other factors with significant correlation are cardiovascular disease prevalence and diabetes mellitus, having least cluster means: **-0.189, −0.212** respectively. The results show second highest percentage of health expenditure (**-0.412**) and PM_2.5_ level (**-0.351**).

**Table 4.** Cluster Mean of Variables for COVID-19 Confirmed Deaths

| Feature Variables | Cluster1 | Cluster2 | Cluster3 | Cluster4 |
| --- | --- | --- | --- | --- |
| Cum_deaths | 0.241 | -3.06e-05 | 1.260 | -0.330 |
| GDP_per_capita | 1.367 | -0.3008 | -0.611 | -0.763 |
| EPI | 1.371 | -0.079 | -1.187 | -1.035 |
| HLT | 1.451 | -0.100 | -0.957 | -1.097 |
| AIR | 1.475 | -0.181 | -1.184 | -0.983 |
| PMD | 1.392 | -0.351 | -0.996 | -0.680 |
| HAD | 1.220 | 0.043 | -1.007 | -1.088 |
| H<sub>2</sub>O | 1.344 | 0.00016 | -0.688 | -1.167 |
| UWD | 1.412 | -0.090 | -0.633 | -1.105 |
| Asthma | -0.084 | -0.246 | 5.277 | -0.027 |
| Cardiovascular | -0.0219 | -0.189 | 5.494 | -0.185 |
| Diabetes Mellitus | -0.0823 | -0.212 | 5.7142 | -0.1162 |
| Nutritional Def. | -0.218 | -0.1891 | 5.172 | 0.0212 |
| Respiratory Infect. | -0.208 | -0.194 | 6.043 | -0.0561 |
| Health exp | 1.441 | -0.412 | -0.615 | -0.670 |
| tuberculosis cases | -0.2794 | -0.250 | 4.956 | 0.183 |
| Alcohol consumption | 0.578 | 0.128 | 0.046 | -0.7155 |
| Smoking prevalence | 0.142 | 0.203 | -0.329 | -0.393 |
| Life expectancy | 1.122 | 0.118 | -0.400 | -1.160 |

#### 5.2.3. Analysis of Cluster 3

This cluster only comprises of China and India as shown in Table 2c. It has the highest mean of COVID-19 death cases (**1.260**) as visible from cluster means in Table 4. This cluster also has the highest mean of COVID-19 confirmed cases. From the correlation matrix, cumulative death cases are significantly positively correlated with asthma prevalence (**0.5**), cardiovascular disease prevalence (**0.4**), diabetes mellitus (**0.4**), nutritional deficiencies (**0.4**), respiratory infections (**0.3**) and tuberculosis cases (**0.3**). This cluster also shows the highest cluster means (**5.277, 5.494, 5.714, 5.172, 6.043, 4.95**) of these factors respectively as compared to other clusters.

India has disease prevalence of respiratory infections, cardiovascular and diabetes. While, cardiovascular diseases, chronic obstructive pulmonary diseases (COPD) and diabetes are the leading causes of deaths in China. Higher rate of Chronic obstructive pulmonary disease (COPD) in China is mainly due to the effective diagnosis and preventive measures to help increase life expectancy [51]. There are many secondary afflictions linked with COPD such as cardiovascular disease, diabetes mellitus, osteoporosis and anxiety. People with such conditions are more vulnerable to COVID-19. Air pollution and dietary risks are other major risk factors for higher death rates in China.

#### 5.2.4. Analysis of Cluster 4

This cluster shows least cluster mean of confirmed deaths **(−0.330**). Cluster means (%) of asthma prevalence (**-0.027**), cardiovascular disease prevalence (**-0.185**), diabetes mellitus (**-0.116**), nutritional deficiencies (**0.021**), respiratory infections (**-0.056**) and tuberculosis cases (**0.18**) show second highest cluster mean. Countries in this cluster are developing countries such as Afghanistan, Nepal, Sudan, Djibouti, Ethiopia, Mauritania, Madagascar and Senegal. They have limited healthcare facilities such as hospitals and clinics. Nepal and Tanzania lack hand-washing stations, appropriate garbage removal systems, running water and germ-free medical equipment, resulting in transmission of diseases. On the contrary, these countries show low COVID-19 death rates as compared to other clusters. Undoubtedly, these countries do not have sufficient testing facilities but there can be other reasons of low reported death cases such as Senegal have faced the major outbreak of Ebola epidemic. Its experience has lead to better preparation, and a well timed authorities’ response resulting in less number COVID-19 cases [52].

## 6. Visualization of Clusters

Choropleth maps are used for visualization of results. Maps are generated for the available data of 79 countries to visualize the results of K-Means for COVID-19 confirmed cases and COVID-19 death cases in different countries. First plot in Fig. 4a shows countries on the basis of COVID-19 confirmed cases. Annotations represent United States having 15,648,098 cases as maximum count. Fig. 4b shows the clustering of countries on the basis of K-Means. The visualization makes it easier to understand the grouping of countries on the basis of related factors. Fig. 4b shows that there are 4 clusters in total. China and India are the only two countries in cluster 2 (cluster 3 in Table. 2c). Cluster 0 (cluster 1 in Table. 2a) is the largest and a detailed correlation analysis has been provided for each cluster in Section 4 and Section 5. Fig. 5a shows COVID-19 deaths in 79 countries across the world. Highest number of deaths are in USA and the color shades show different groups of countries on the basis of death cases. Fig. 5b shows the clusters formed on the basis of K-Means considering socio-economic factors, disease prevalence and other environmental health indicators and cumulative death cases.

**Figure 4:**
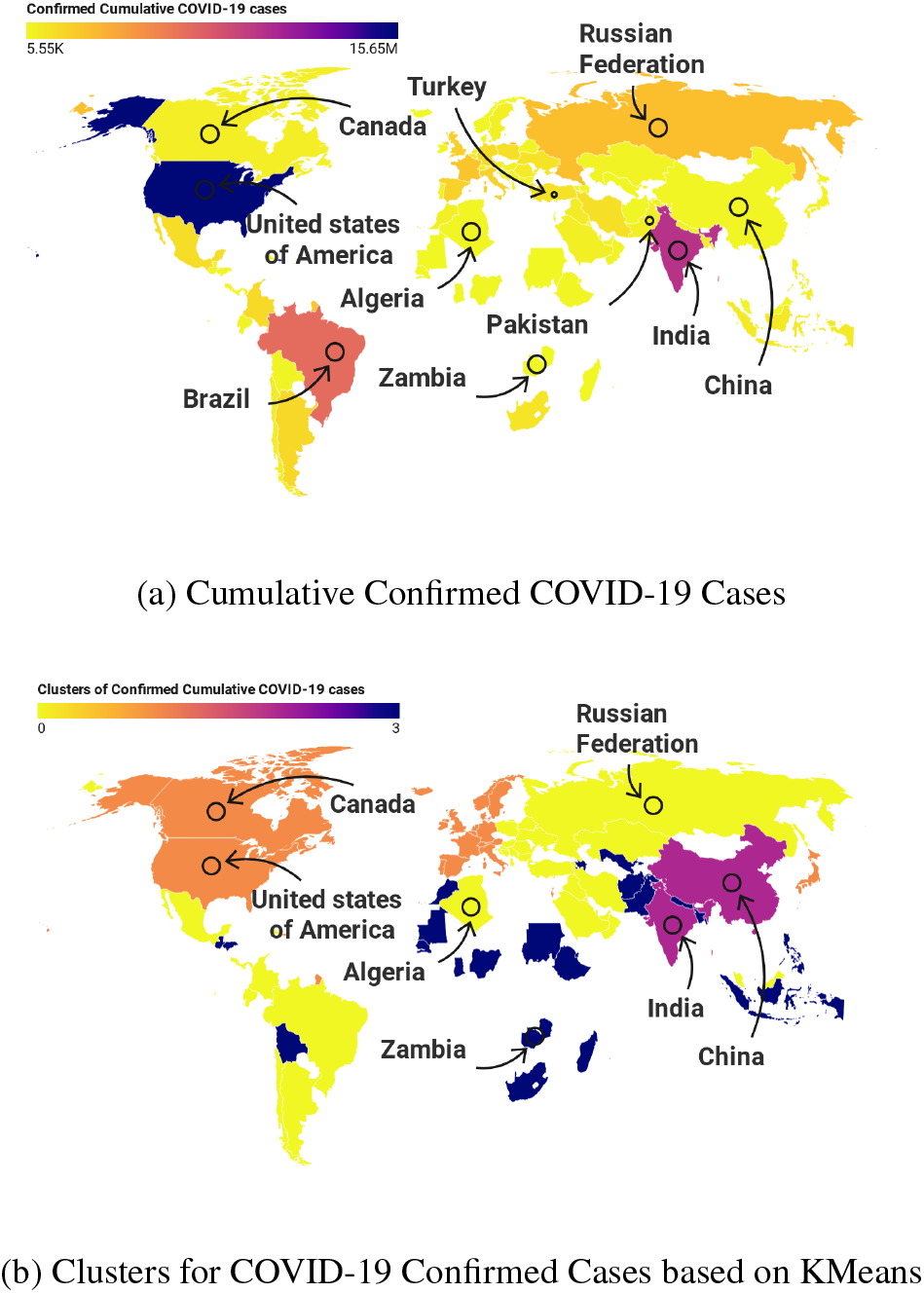
Choropleth Maps for COVID-19 Confirmed Cases

**Figure 5:**
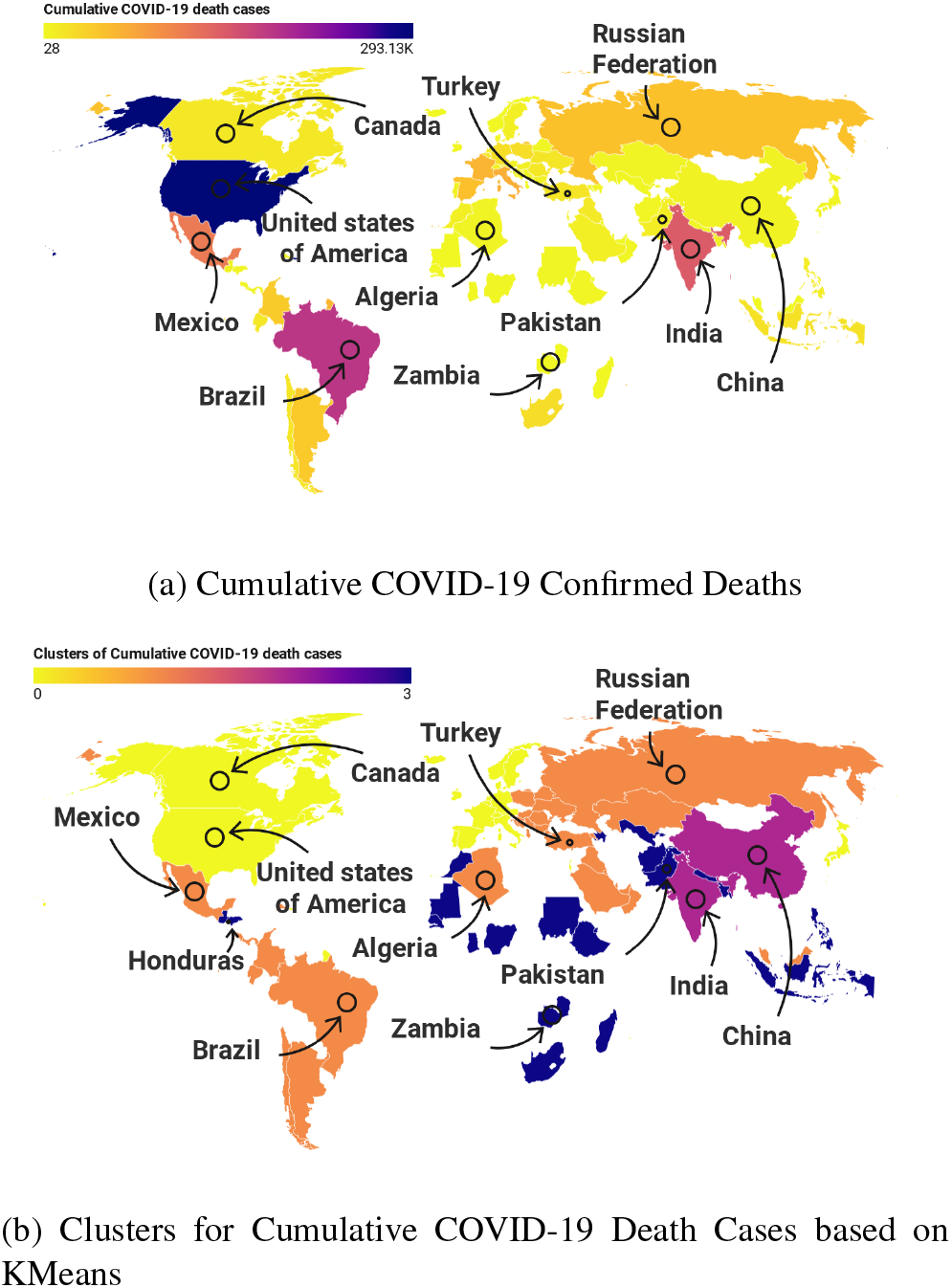
Choropleth Maps for COVID-19 Death Cases

## 7. Conclusion

Unsupervised K-Means algorithm is used in this paper to cluster 79 countries on the basis of socio-economic, disease prevalence and health system indicators considering COVID-19 confirmed cases and COVID-19 death cases as evaluation parameters in order to study the factors closely involved in the spread of disease. Elbow method is used to find the optimal number of clusters. Asthma prevalence, diabetes mellitus, cardiovascular disease prevalence, nutritional deficiencies and health expenditure show significant positive correlation with cumulative COVID-19 confirmed cases. Four clusters are formed applying K-Means on COVID-19 confirmed cases and COVID-19 death cases. Cluster 1 consists of 33 countries with developed as well as developing countries showing third highest cluster mean percentage of COVID-19 confirmed cases and COVID-19 death cases. Cluster 2 contains developed countries with second highest cluster mean percentage for COVID-19 confirmed cases and death cases. Cluster 3 consists of only two countries: China and India, showing highest cluster mean percentage of COVID-19 confirmed cases and COVID-19 death cases. Cluster 4 contains 23 developing countries, and has least cluster mean percentage of COVID-19 confirmed cases and COVID-19 death cases. Disease prevalence are strongly associated with COVID-19 while environmental health indicators are weakly associated with COVID-19. The results produced can be utilized by policy makers to make betters decisions to control the pandemic.

## Data Availability

All data sources have been mentioned in the manuscript.

---

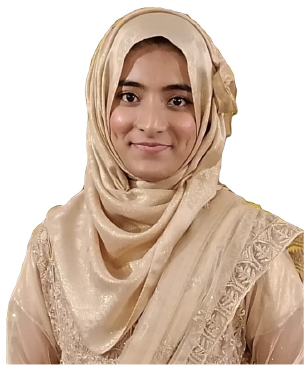

Syeda Amna Rizvi is a postgraduate student at University of Engineering and Technology, Lahore. She received her BSc degree in Computer Engineering from University of Engineering and Technology, Taxila in 2019. Her research interests include Machine Learning, Data Sciences, Exploratory and Multivariate Data Analysis, Data Visualization and Data Wrangling.

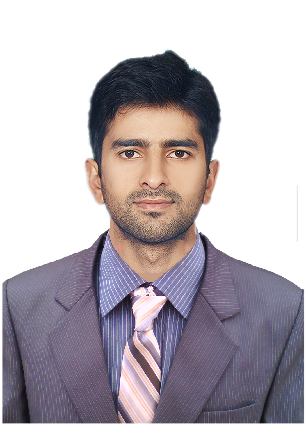

Muhammad Umair is a Lecturer at Department of Electrical, Electronics and Telecommunication Engineering, New Campus, UET Lahore. He completed his B.Sc. Electrical Engineering and M.Sc. Electrical Engineering from University of Engineering & Technology (UET) Lahore in 2014 and 2017, respectively. He has worked as a Research Officer at Internet of Things (IoT) lab at Al-Khwarizmi Institute of Computer Sciences, UET Lahore. He has also worked at Sultan Qaboos IT Research lab as a Research Officer. His areas of interests include Internet of Things, Embedded Systems, Network Systems, Machine Learning, Algorithms Development, Ubiquitous Computing, Cloud Based Systems, Data Analytics and working on application layer of any of the defined problems.

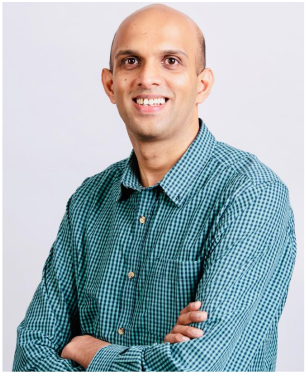

Muhammad Aamir Cheema is an ARC Future Fellow, an Associate Professor and Director of Research at the Department of Software Systems and Cybersecurity, Faculty of Information Technology, Monash University, Australia. He obtained his PhD from UNSW Australia in 2011. He is the recipient of 2012 Malcolm Chaikin Prize for Research Excellence in Engineering, 2013 Discovery Early Career Researcher Award, 2014 Dean’s Award for Excellence in Research by an Early Career Researcher, 2018 Future Fellowship, 2018 Monash Student Association Teaching Award and 2019 Young Tall Poppy Science Award. He has also won two CiSRA best research paper of the year awards, two invited papers in the special issue of IEEE TKDE on the best papers of ICDE, and three best paper awards at ICAPS 2020, WISE 2013 and ADC 2010, respectively. He is the Associate Editor of IEEE TKDE and DAPD and served as PC co-chair for ADC 2015, ADC 2016, 8th ACM SIGSPATIAL Workshop ISA 2016 & 2018, IWSC 2017, proceedings chair for DASFAA 2015 & ICDE 2019.

